# Effects of virtual reality-based aerobic endurance training on the functional fitness of healthy older adults: A systematic review

**DOI:** 10.1101/2022.05.24.22275497

**Authors:** Daniela Ramírez Restrepo, Julialba Castellanos Ruiz, Lina María Montealegre Mesa, Carolina Márquez Narváez, Santiago Murillo Rendón

## Abstract

**Objetive:** to analytically and systematically review and integrate the available evidence of the effects of virtual reality-based aerobic endurance training on functional fitness of healthy older adults.

**Materials and Methods:** This systematic review of randomized controlled trials was conducted through searches in Pubmed, Science Direct, Scopus, PEDro, Web of Science, Lilacs, Scielo, Dialnet, and IEEE Xplore between February and May 2021. The evaluation of bias and methodological quality was performed following the parameters proposed by the Cochrane Manual of Systematic Reviews of Interventions and the PEDro Scale. Review Manager Software (Version 5.4.1) served for a summary of bias.

**Results:** Three clinical trials were selected. None of the selected studies employed any exclusively aerobic intervention with virtual reality; however, statistically significant findings for this type of intervention were found in functional fitness such as aerobic endurance, upper and lower limb strength, agility, and dynamic balance.

**Conclusions:** aerobic endurance training through the use of virtual reality may provide positive effects on the functional fitness of the healthy elderly. This has implications for clinical practice, as it fosters the development of more beneficial, attractive, motivational, and quality interventions, the prevention of common conditions, and the use of technological tools by the elderly population. However, as for research implications, an increase of knowledge on the effects of technologies in the elderly functional capacity, and an implementation of scientific models or theories on physical activity and exercise are recommended with the purpose of explaining the phenomena due to the use of technologies within the processes of functional rehabilitation.

## Introduction

The demographic and epidemiological transitions during the last decades have brought about a significant global increase in the number of people aged 60 years and older.(1) While in 2000 the percentage of elderly population represented 10% of the total population, it is estimated that by the year 2030 it will represent 16.6% of the total population and 22% by the year 2050.(2) A longer life expectancy makes individuals, particularly the elderly, prone to bodily impairments, functional limitations and restrictions in social participation. This is due to the fact that, in old age, there may be a greater number of health, environmental, and socioeconomic risks, as a result of a longer life trajectory.(3) Additionally, determinants such as unhealthy lifestyles, along with the physiological changes during the aging process may increase the probability of different health conditions that involve their physical activity such as aerobic endurance.(4–6)

Aerobic endurance is a remarkable component of functional fitness throughout the lifetime, as it allows for the independent performance of daily life, instrumental, and social activities with a lower level of fatigue.(7) This physical activity tends to decrease in older adults due to the cardiovascular and pulmonary diseases during the aging process.(8,9) Therefore, reduced cardiac contractility, conduction velocity and baroreceptor sensitivity, decreased cardiac output and blood flow, valvular insufficiency, increased arterial wall stiffness, decreased thoracic, alveolar and pulmonary distensibility, loss of valvular and respiratory muscle strength, reduced cilia and cough reflex, among others, can reduce pulse rate during exercise. Additionally they can reduce cardiac reserve, and increase blood pressure. Reduce loss of strength of valvular and respiratory muscles, reduction of cilia and cough reflex, among others, can decrease pulse rate during exercise, reduce cardiac reserve, increase blood pressure, decrease gas exchange surface area and increase the probability of heart failure, arrhythmias, other cardiovascular diseases, infections or respiratory diseases in old age.(10–12) Also, these changes may significantly affect functional independence in the aforementioned daily life activities.(7)

Therefore, aerobic fitness is one of the most recommended strategies to maintain or improve functional fitness in this population.(13,14) Leirós et al., for example, demonstrated that aerobic training for the elderly markedly improved functional fitness such as strength and flexibility of upper and lower limbs, arm strength, and weight loss and reduced BMI.(15) Similarly, Ribeiro et al. found that water-based aerobic training showed positive findings in functionality, as well as in some aspects related to quality of life in women over 65 years of age.(16)

Great technological advances have allowed for an improvement in the effects and quality of health care, diagnosis, treatment, and intervention strategies.(17,18) For functional rehabilitation in the elderly, the incorporation of high technologies for functional fitness training, as virtual reality, has brought a lot of benefits. Performing activities in virtual environments, that resemble the real ones, and the visual and auditory feedback provided by these tools, increase individuals’ motivation to exercise.(19–21) According to the Ecological Model of Physical Activity, it has been established that, beyond the influence of biological factors such as fitness level, health status, or body composition, there are psychological and environmental factors that influence and enhance the physiological effects of exercise.(22–24)

Although some benefits of conventional aerobic training and some advantages of virtual reality are already known, few studies have been conducted on the effects of this type of intervention on the functional fitness of older people, especially those with stable or healthy conditions. Van der Kolk et al., for example, evaluated the effectiveness of home and remote training in people between 30 and 75 years of age with Parkinson’s Disease, which included the use of serious games (exergames) through Virtual Reality software. As results, the authors found improvements in the motor status of the participants in the experimental group, in contrast to the control group.^(25)^ Similarly, Karssemeijer et al. evaluated the efficacy of training with exergames and conventional aerobic training in frail older people with dementia. There, it was found that the intervention with these serious games, compared to the people in the control group, yielded positive results in reducing the levels of fragility in the participants of the experimental group.^(26)^

Thus, the objective of the present study was to analytically and systematically review and integrate the available evidence of the effects of virtual reality-based aerobic endurance training on functional fitness of healthy older adults. It is expected that the collection of information recorded in this research will serve as a point of reference for future studies in gerontological physiotherapy, especially to promote the implementation of technological devices in favor of the improvement of conventional interventions, as well as functionality, quality of life, and social participation in old age. This research aims to respond to these realities such as digital evolution, global demographic aging and stereotypes associated with the inability of older people regarding the use of technologies ^(1,27,28)^ by encouraging the transition from traditional schemes of functional rehabilitation in old age toward schemes of higher quality and demystifying existing social stigmas regarding aging, old age, and the use of technological tools.

## Material and Methods

### Type of Study

This systematic review of Randomized Controlled Trials (RCTs) was conducted on the effects of aerobic resistance training through virtual reality on the functional physical condition of healthy older people. The PICO strategy was used to guide the formulation of the research question, where P (patient or target population) = Healthy Older People; I (intervention) = Aerobic Endurance Training through Virtual Reality, either exclusively or combined with the training of other physical capacities; C (comparison or control) = Conventional Aerobic Endurance Training or No Intervention; OR (outcomes) = Aerobic Endurance (primary outcome measure), Muscular Strength, Flexibility and Balance (secondary outcome measures). Subsequently, the following research question established: What are the effects of aerobic endurance training through virtual reality on the functional physical condition of healthy older adults?

### Search Strategy

Searches were conducted in PubMed, Science Direct, Scopus, PEDro, Web of Science, Lilacs, Scielo, Dialnet, and IEEE Xplore databases. The search was executed up to the month of May 2021 and search equations were applied that combined the following MeSH terms to identify relevant studies: Aged, Virtual Reality, Aerobic Exercise, Physical Fitness. Likewise, related terms such as Elderly, Virtual Reality Exposure Therapy, Aerobic Exercises, Aerobic Training were used. For databases such as Lilacs and Dialnet, searches were also carried out with equations in the Spanish language, using DeCS terms such as Virtual Reality, Aerobic Training, Functional Physical Performance, and Elderly. All terms described were combined with the Boolean operators “AND” and “OR” to perform multiple combinations and generate several search equations. Finally, the references of some scientific articles were reviewed to identify if there were relevant studies for the present research.

### Study Inclusion and Exclusion Criteria

The study selection comprised RCTs in any language, of healthy or with any controlled chronic health condition people of 60 years or older and that were conducted during the last 20 years (2001-2021). Other types of studies were excluded, as well as articles of people with neurological, psychiatric, or mental health conditions within their target population.

### Procedures

#### Study Selection

Searches mentioned above in databases were carried out by one researcher, using the established search equations. First, articles were reviewed according to their titles and abstracts; then, the full text analysis of the articles that met the inclusion and exclusion criteria of this research was carried out.

#### Extraction and Management Data

The required information was extracted by a researcher. To register the articles of interest, tables were created in the Review Manager Software (Version 5.4.1)^(29)^ that included data such as the name of the study, authors, year of publication, methods (type of study, method of intervention allocation, groups, loss to follow-up), participants (location, time frame of the study or intervention, inclusion and exclusion criteria, sociodemographic characteristics of the population), prescription parameters of the interventions of the control groups and experimental (exercise type/mode, frequency, duration, intensity), and results.

#### Assessment of the Bias Risk and Methodological Quality of the Articles Included

Evaluation of bias and methodological quality were performed following the parameters proposed by the Cochrane Manual of Systematic Reviews of Interventions and the PEDro Scale.^(30,31)^ First, selection, performance, detection, attrition, and reporting biases were considered; this in order to determine, respectively, the systematic differences between: 1) the characteristics of the comparison groups at the beginning of the study; 2) experimental and control groups for the intervention of interest; 3) the way in which the results between the experimental group and the control group were determined; 4) groups regarding study dropouts; and 5) the findings of the study, both presented and not presented. Similarly, other biases associated with problems that were not considered in the previous types of biases were taken into account.^(30)^

Based on the biases described above, each type of bias in the included studies was assessed, classifying each as low bias, high bias, or unclear bias^(30)^ Second, two researchers independently evaluated the methodological quality of the studies included through 11 items of the PEDro Scale, which presented dichotomous options as alternatives response (If: 1 point. No: 0 points) These items were^**(31)**^:

1. The selection criteria were specified
2. Subjects were randomly assigned to groups (in a crossover study, subjects were randomly distributed as they received treatments).
3. It was a blinded allocation
4. The groups were similar at baseline in relation to the most important prognostic indicators.
5. All individuals were blinded for treatment.
6. All therapists were blinded for treatment administration.
7. All evaluators were blinded for measuring at least one key outcome.
8. Measures of at least one of the key outcomes were obtained from more than 85% of the subjects who were initially assigned to the groups.
9. Results were presented for all subjects who received treatment or were assigned to the control group; on the contrary case, data for at least one key outcome were analyzed on an “intention-to-treat” basis.
10. Results of statistical comparisons between groups were reported for at least one key outcome.
11. This study shows point and variability measures for at least one key outcome.

Finally, these evaluations were socialized together to reach a consensus and, in cases of disagreements, these were discussed and resolved with a third evaluator. Similarly, the Cochrane’s Review Manager (Version 5.4.1) was used for the creation of the summary bias analysis.(25)

## Results

### Study Selection

A total of 241 articles from databases were selected. Then, 74 duplicate articles were removed, for a total of 167 screening articles. Out of the 167, 164 were excluded based on title, abstract and inclusion and exclusion criteria, resulting in a final number of three articles for full-text review and qualitative assessment. The details of the study selection are depicted in the PRISMA flow chart (Fig. 1)

**Figure 1.**
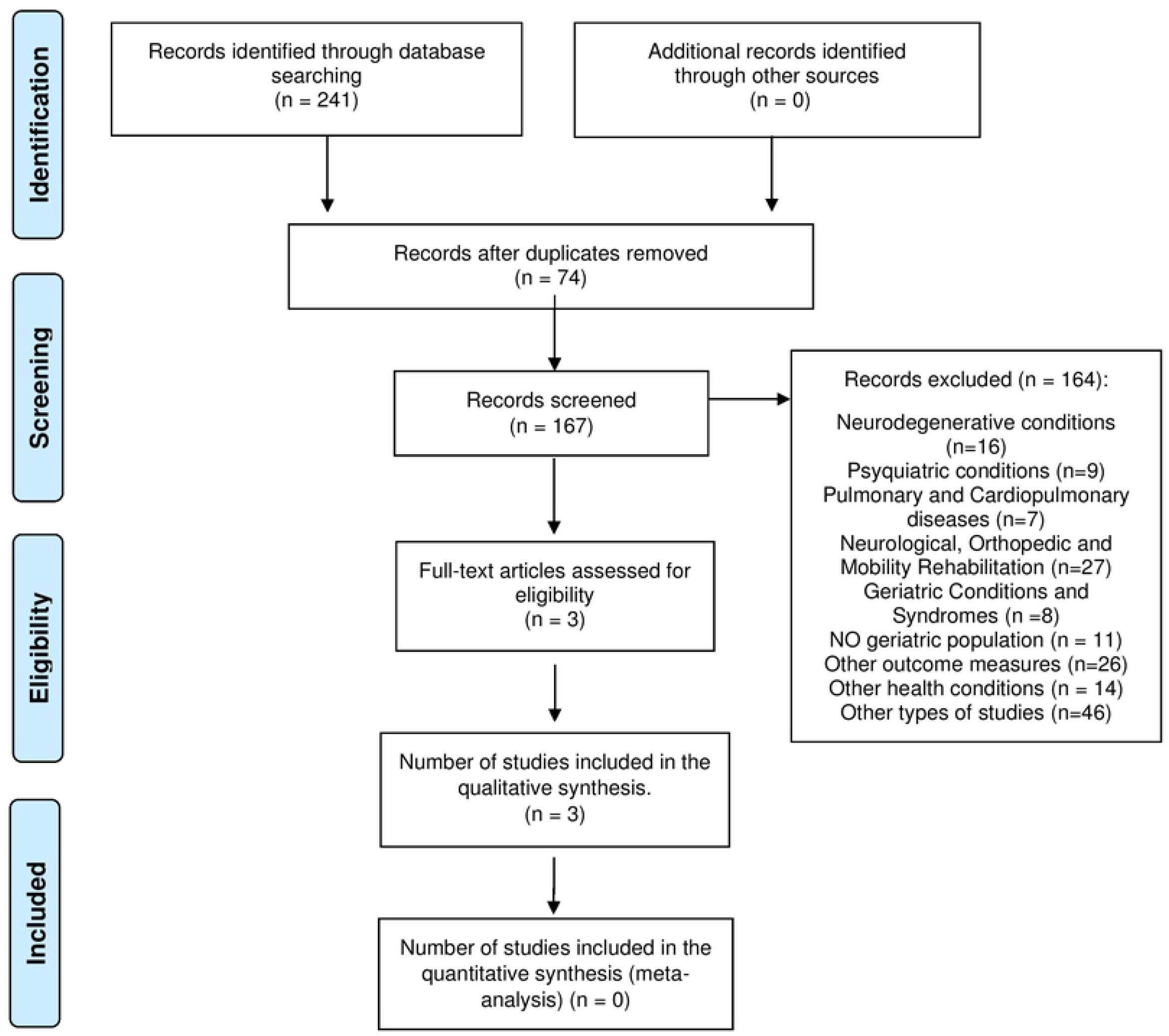
PRISMA flow diagram.

### Characteristics of the Selected Studies

Table 1 shows the characteristics of selected studies. This systematic review comprised randomized controlled trials that measure the effects of aerobic endurance training through the use of virtual reality on the functional fitness in the healthy elderly in comparison with traditional training or absence of therapeutic intervention. Two of the studies made exclusive use of virtual reality and one of augmented reality to intervene with the experimental group; although, none of them intervened with the control group through the use of conventional therapies or exercises. Similarly, none of the selected studies involved a unique intervention for aerobic capacity in the target populations. Most of the studies were conducted on the Asian continent and their samples included a larger number of women than men.

**Table 1.**
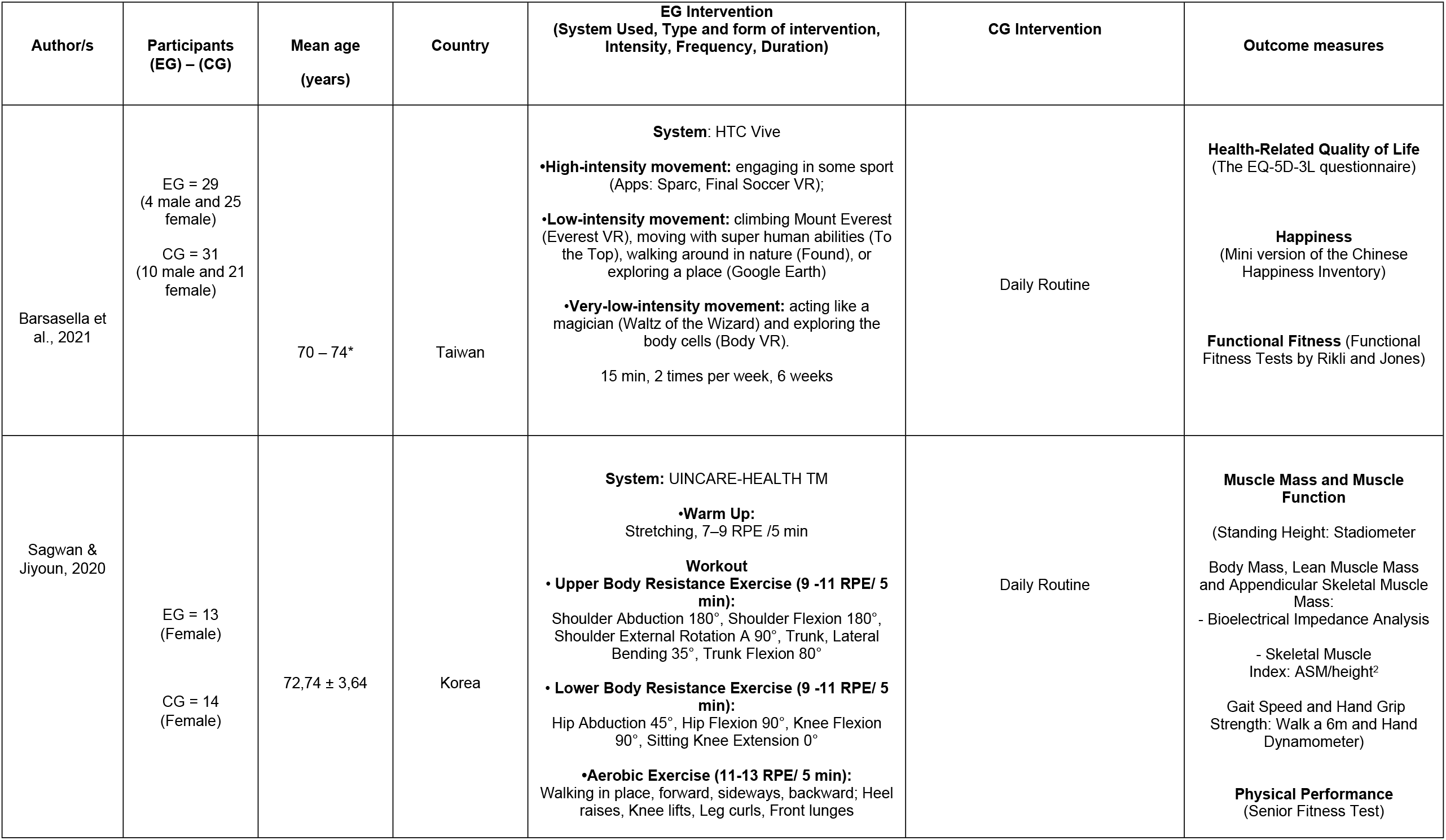
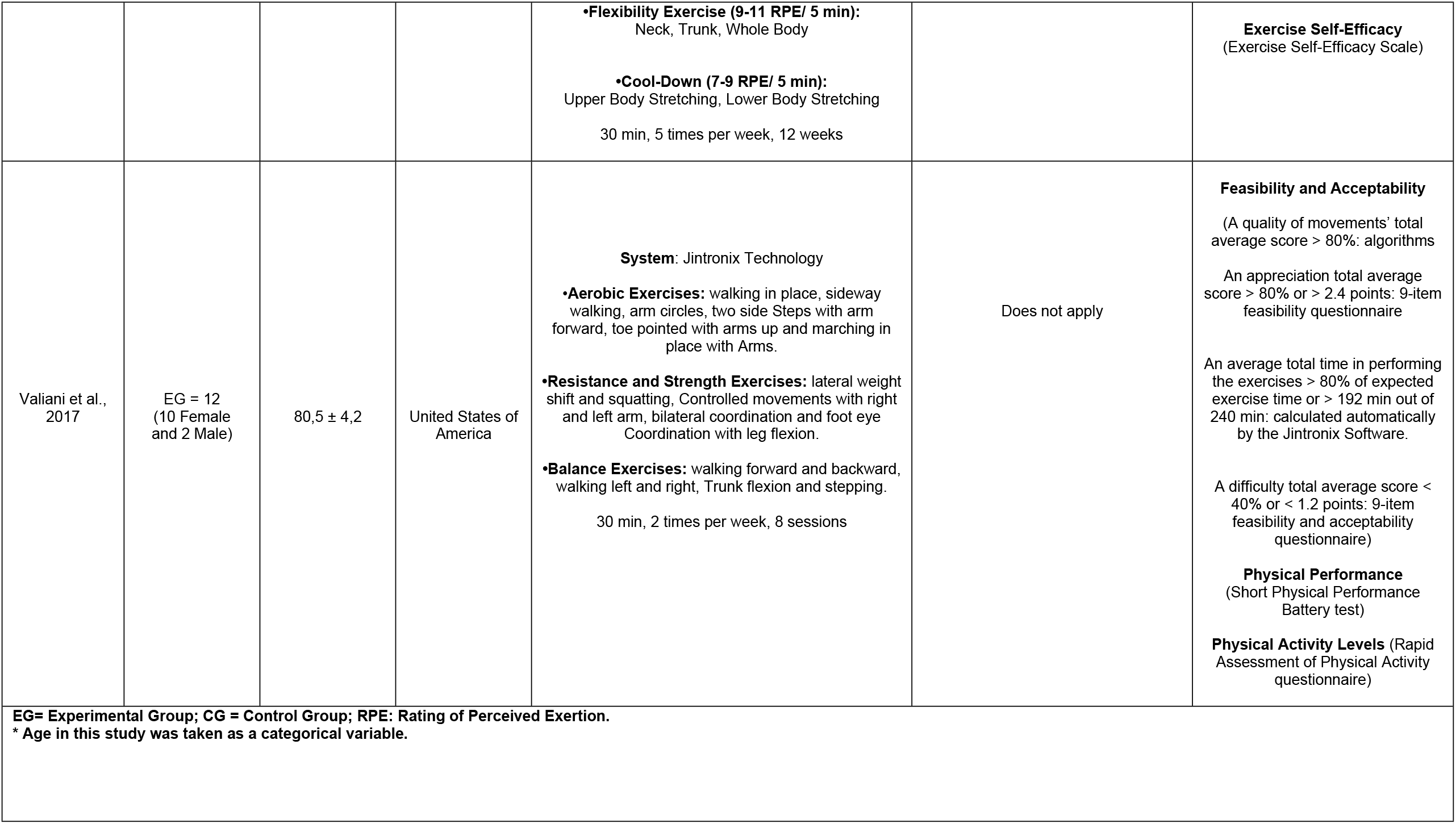
Characteristics of the Selected Studies.

| Author/s | Participants<br>(EG) – (CG) | Mean age<br>(years) | Country | EG Intervention<br>(System Used, Type and form of intervention,<br>Intensity, Frequency, Duration) | CG Intervention | Outcome measures |
| --- | --- | --- | --- | --- | --- | --- |
| Barsasella et al., 2021 | EG = 29<br>(4 male and 25 female)<br>CG = 31<br>(10 male and 21 female) | 70 – 74* | Taiwan | **System:** HTC Vive<br>•**High-intensity movement:** engaging in some sport (Apps: Sparc, Final Soccer VR);<br>•**Low-intensity movement:** climbing Mount Everest (Everest VR), moving with super human abilities (To the Top), walking around in nature (Found), or exploring a place (Google Earth)<br>•**Very-low-intensity movement:** acting like a magician (Waltz of the Wizard) and exploring the body cells (Body VR).<br>15 min, 2 times per week, 6 weeks | Daily Routine | **Health-Related Quality of Life**<br>(The EQ-5D-3L questionnaire)<br>**Happiness**<br>(Mini version of the Chinese Happiness Inventory)<br>**Functional Fitness** (Functional Fitness Tests by Rikli and Jones) |
| Sagwan & Jiyoun, 2020 | EG = 13<br>(Female)<br>CG = 14<br>(Female) | 72,74 ± 3,64 | Korea | **System:** UINCARE-HEALTH TM<br>•**Warm Up:**<br>Stretching, 7–9 RPE /5 min<br>**Workout**<br>• **Upper Body Resistance Exercise (9 -11 RPE/ 5 min):**<br>Shoulder Abduction 180°, Shoulder Flexion 180°, Shoulder External Rotation A 90°, Trunk, Lateral Bending 35°, Trunk Flexion 80°<br>• **Lower Body Resistance Exercise (9 -11 RPE/ 5 min):**<br>Hip Abduction 45°, Hip Flexion 90°, Knee Flexion 90°, Sitting Knee Extension 0°<br>•**Aerobic Exercise (11-13 RPE/ 5 min):**<br>Walking in place, forward, sideways, backward; Heel raises, Knee lifts, Leg curls, Front lunges | Daily Routine | **Muscle Mass and Muscle Function**<br>(Standing Height: Stadiometer<br>Body Mass, Lean Muscle Mass and Appendicular Skeletal Muscle Mass:<br>- Bioelectrical Impedance Analysis<br>- Skeletal Muscle Index: ASM/height<sup>2</sup><br>Gait Speed and Hand Grip Strength: Walk a 6m and Hand Dynamometer)<br>**Physical Performance**<br>(Senior Fitness Test) |
|  |  |  |  | •**Flexibility Exercise (9-11 RPE/ 5 min):**<br>Neck, Trunk, Whole Body<br>•**Cool-Down (7-9 RPE/ 5 min):**<br>Upper Body Stretching, Lower Body Stretching<br>30 min, 5 times per week, 12 weeks |  | **Exercise Self-Efficacy**<br>(Exercise Self-Efficacy Scale) |
| Valiani et al., 2017 | EG = 12<br>(10 Female and 2 Male) | 80,5 ± 4,2 | United States of America | **System:** Jintronix Technology<br>•**Aerobic Exercises:** walking in place, sideway walking, arm circles, two side Steps with arm forward, toe pointed with arms up and marching in place with Arms.<br>•**Resistance and Strength Exercises:** lateral weight shift and squatting, Controlled movements with right and left arm, bilateral coordination and foot eye Coordination with leg flexion.<br>•**Balance Exercises:** walking forward and backward, walking left and right, Trunk flexion and stepping.<br>30 min, 2 times per week, 8 sessions | Does not apply | **Feasibility and Acceptability**<br>(A quality of movements' total average score > 80%: algorithms<br>An appreciation total average score > 80% or > 2.4 points: 9-item feasibility questionnaire<br>An average total time in performing the exercises > 80% of expected exercise time or > 192 min out of 240 min: calculated automatically by the Jintronix Software.<br>A difficulty total average score < 40% or < 1.2 points: 9-item feasibility and acceptability questionnaire)<br>**Physical Performance**<br>(Short Physical Performance Battery test)<br>**Physical Activity Levels** (Rapid Assessment of Physical Activity questionnaire) |

EG= Experimental Group; CG = Control Group; RPE: Rating of Perceived Exertion.
\* Age in this study was taken as a categorical variable.

### Analysis and Synthesis of Results

Due to the diversity of the selected studies in terms of sample size, intervention procedure, and outcome measures, performing a quantitative analysis (meta-analysis) was not possible. However, a qualitative analysis showed positive results regarding the effects of aerobic training through the use of virtual reality on the functional fitness of the healthy elderly.

Regarding aerobic endurance, only one of the studies showed statistically significant improvements, after an augmented reality intervention, in the two-minute walk test of the Senior Fitness Test for the experimental group (p=0.020).(26) However, for the other activities that make up functional fitness, this study also showed positive findings in muscle function (p=0.003) and lower limb muscle strength (p=0.033), as well as in agility and dynamic balance (p=0.032) of the intervention group.(26)

Similarly, another selected study exhibited a statistically significant improvement in upper limb strength, agility and dynamic balance for the experimental group after the virtual reality intervention (p≤0.05).(27) Also, the third selected showed significant improvements in the gait speed test (p≤0.05) and in the test of getting up and sitting in a chair (p≤0.05), after the intervention of the experimental group by using virtual environments.^(34)^ With respect to this last study, although the walking speed and the sitting and standing up from a chair test are specific tasks, these require functional fitness such as muscular strength, mainly in the lower limbs, aerobic capacity and static and dynamic balance.(28,29)

### Risk of Bias in Included Studies

The summary on the risk of bias assessment is in Figure 2. One of the studies was judged to have an unclear risk of selection bias since, although an experimental group was established, detailed information on the sequence generation process and concealment is not provided.(30) Additionally, an unclear risk of performance bias was determined in the same study as information on blinding of participants and personnel was insufficient.(30)

**Figure 2.**
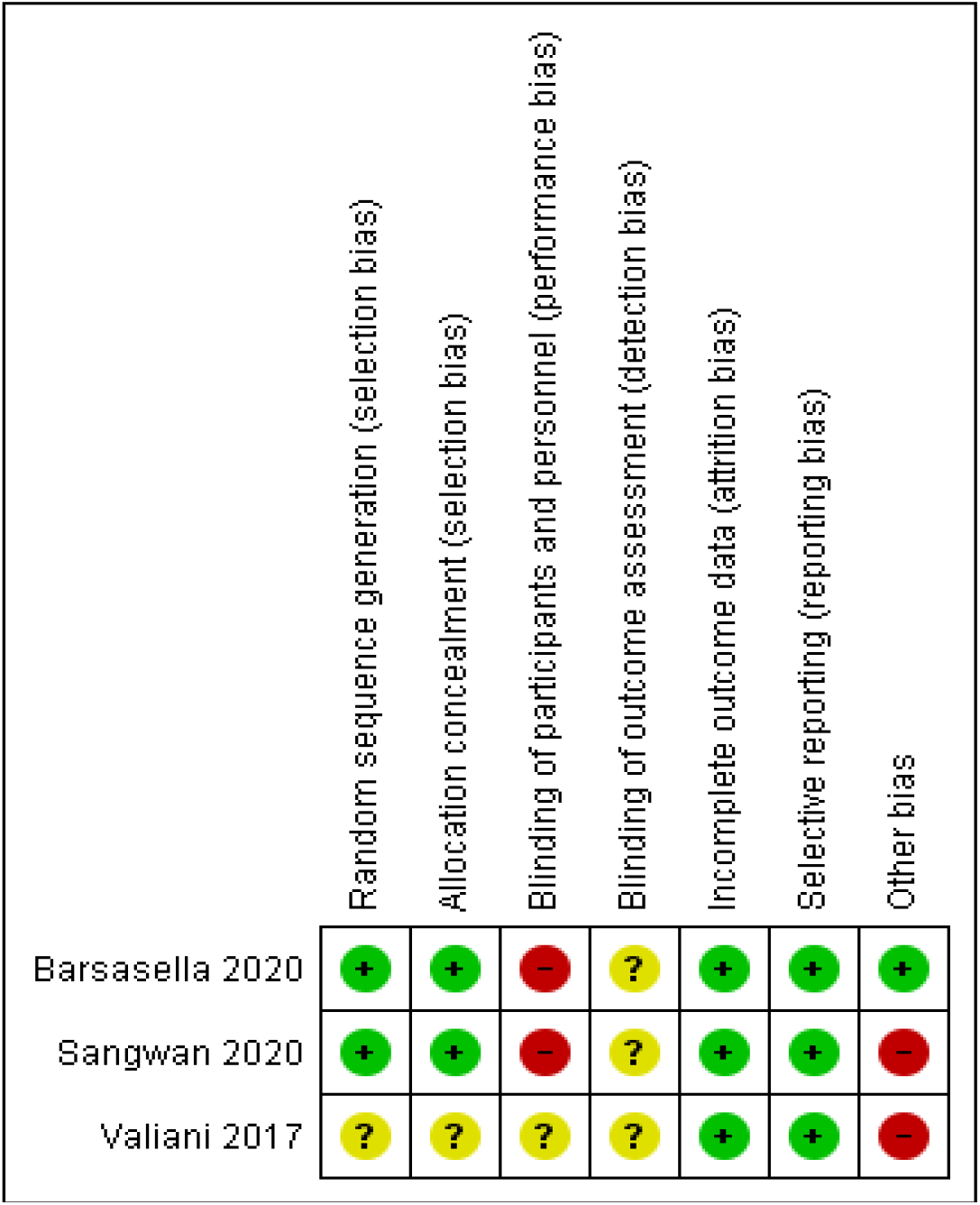
Summary Risk of Bias in Included Studies.

Regarding detection bias, all included studies were rated at unclear risk, since none of them addressed the results about blinding of assessors to their research findings.(26,27,30) However, all studies were judged to be at low risk of attrition and reporting bias as missing outcome data were not presented and the protocol and results for each study were explicitly stated.(26,27,30) Finally, in relation to other types of bias, two studies were considered high risk since the method of assigning the experimental group was not specified in any of them(30) and the type of study was not described in the methodology of the remaining one.(26)

## Discussion

This systematic review investigated the effects of aerobic endurance training through virtual reality on the functional physical condition of healthy older people based on the findings of three RCTs. The results suggest that the interventions, although they were not exclusively aerobic, have a positive influence on physical capacities such as aerobic endurance, muscular strength, agility and dynamic balance in people aged 60 or 65 and older.

First, although few studies were included, the findings of this review are consistent with the results of certain research available within the scientific evidence. The positive findings of the Barsasella study regarding aerobic capacity(27) are consistent with the results of another investigation, which found that its experimental group, after the implementation of an exercise program with virtual reality, reached maximum heart rate and metabolic cost in a shorter period of time and significantly. This is attributed, according to this research, to the fact that immersion experiences, simulated environments, life-like experiences and visual feedback could possibly make it easier for participants to tolerate higher intensities of exercise and perceive less sensation of actual physical discomfort.(31)

Similarly, some of the findings of the study by Valiani et al., as well as the study(30) by Barsasella,(27) coincide with the results of other studies,(32,33) in which improvements in capacities such as aerobic endurance, appendicular muscle strength, flexibility, and dynamic balance were also found after interventions with reality virtual. The results of these investigations, according to their authors, were probably presented thanks to the level of attraction and security generated by virtual environments, since these can facilitate adherence to exercise and continuity in its execution by the target populations.(32,33)

Secondly, from the perspective offered by the Ecological Model of Physical Activity, one may also think that the findings of the studies included in this review are due to the benefits offered by virtual reality. In this sense, it is likely that extrinsic factors such as feedback, the ecological tasks provided to the user and the simulation of real environments offered by this type of technology, stimulate a series of psychological factors such as motivation, pleasure, fun, revitalization, and a sense of tranquility, achieving that the physiological effects of the types of exercises (which are purely biological factors), are enhanced and produce more significant improvements in the functional physical condition of the elderly than conventional training programs.(23,31,33,34) Moreover, extrinsic and psychological factors that are stimulated by the use of technologies such as virtual reality can provide multiple ways to perform activities and facilitate the adherence and establishment of exercise as a habit in this population. These advantages, in the end, are often more difficult to achieve or realize when working in the traditional way.(33)

Third, although it was found that the virtual reality interventions employed in the included studies were not designed only for aerobic endurance work, several investigations have shown that this type or mode of exercise is one of the most recommended for improving functional status in old age, as well as for treating or managing chronic conditions(35–39); this is because increased tissue oxygen, increased stroke volume, improvements in perfusion, angiogenesis and increased blood vessel caliber, reduced blood pressure, and increased gas exchange surface area not only promote the function of the cardiovascular and pulmonary systems in the elderly, but also stimulate the synthesis of mitochondria and muscle proteins, improve oxygenation at the cerebral level and stimulate the release of neurotrophic factors.(4,40–42) All of this, ultimately, can translate into an improvement in other physical abilities such as strength, balance and flexibility, as well as cognition. Likewise, in terms of functionality, the maintenance or improvement of functional physical condition can ensure the development of basic, instrumental, and social activities, as well as improve well-being and quality of life during old age.(9)

Finally, this review has strengths and limitations that should be mentioned. Regarding the former, it is worth noting that in this systematic review an exhaustive bibliographic search was carried out in several databases and included studies in all languages, developed over a considerable period of time, to try to collect the most scientific information available regarding the effectiveness of aerobic resistance training by means of virtual reality on the functional physical condition of healthy elderly people. These aspects give relevance and novelty to this research since most of the studies currently developed in the field of physical therapy, aging and old age have investigated the effects of conventional exercise programs in an older adult population with one or more health conditions. However, with respect to the latter, aspects such as the limited evidence available on the topic in question, the absence of a search within the gray literature, the diversity and quality of the included studies, the lack of studies that include exclusive aerobic interventions and the small sample sizes of the RCTs reviewed are highlighted.

## Conclusions

The findings of this review indicate that aerobic endurance training via virtual reality can generate positive effects on functional fitness in healthy older adults. These results have important future implications: for clinical practice, this review demonstrates that interventions that are accompanied by technologies such as virtual reality, whether unimodal or multimodal, can become more beneficial and motivational within the functional rehabilitation processes of individuals aged 60 or 65 years and older, while promoting adherence to exercise and primary or secondary prevention of prevalent health conditions in old age, which represent enormous health costs. In addition, the use of such tools proves to be an attractive, quality and supportive complement, quite useful for health professionals or for interdisciplinary teams that intervene in this population and that will be confronted with the consequences of population aging and the great advances of the technological era.

Similarly, for the elderly, the use of these tools can increase levels of motivation and therapeutic adherence, as well as break down social stereotypes related to their inability to use technology. This is ultimately vital to achieve exercise-related physiological adaptations, produce improvements in function and functionality, achieve a transformation of lifestyle behaviors, increase social participation, intergenerational relationships, and improve well-being and health-related quality of life in old age.

Finally, as implications for research, the results of this study should be carefully evaluated due to the limitations already described. Therefore, it is recommended to increase the generation of knowledge on these topics through experimental studies, which have greater methodological rigor, reduce the risk of bias and allow the generalization of results and recommendations to the population in general. It also reiterates the need to increase research on healthy populations, since prevention strategies during the aging and old age processes are vital for reducing morbidity and mortality rates, the demand for health services, and health costs, among others. Likewise, the review of the theoretical models or theories on physical activity and exercise is suggested, as well as using those that are relevant to explanation with greater scientific rigor, including the phenomena given in light of the research findings.

## Data Availability

All relevant data are within the manuscript and its Supporting Information files.

## Acknowledgements

The authors would like to express their sincere gratitude to the Universidad Autónoma de Manizales and the Ministry of Science, Technology and Innovation (Minciencias) for the support and funding granted to the research project: “Desarrollo de un Sistema de Realidad Virtual para la Implementación de los protocolos de Equilibrio y Fuerza Muscular y de Estabilidad en Adultos Mayores con y sin Problemas de Estabilidad” (Development of a Virtual Reality System for the Implementation of Balance and Muscle Strength and Stability protocols in Older Adults with and without Stability Problems), within which this scientific article arises within the framework of the call 831 of 2021. Likewise, the authors express their gratitude to their research team for their valuable disposition and total admiration for the elderly for being our source of inspiration in our professional endeavors. The authors would like to thank the academic and scientific communities working in the field of health, technology and aging, because without their valuable contributions, continually generating new knowledge that can increasingly benefit the elderly population would not be possible. Finally, they thank Alexandra Suaza Restrepo, who works at the translation center at UAM, for translating this final manuscript, and Gregory Wallace Amos for reviewing it.

## Supporting Information

S1 Table. PRISMA checklist.

## Author Contributions

- **Conceptualization**: Daniela Ramírez.
- **Data curation**: Daniela Ramírez.
- **Formal analysis:** Daniela Ramírez.
- **Methodology:** Daniela Ramírez.
- **Supervision:** Julialba Castellanos, Lina María Montealegre, Carolina Márquez, Santiago Murillo.
- **Validation:** Julialba Castellanos, Lina María Montealegre, Carolina Márquez, Santiago Murillo.
- **Writing – original draft:** Daniela Ramírez.
- **Writing – review & editing:** Julialba Castellanos, Lina María Montealegre, Carolina Márquez, Santiago Murillo.

## Notes

### Competing Interest Statement

The authors have declared no competing interest.

### Funding Statement

The funders had no role in study design, data collection and analysis, decision to publish, or preparation of the manuscript.

### Author Declarations

Universidad Autónoma de Manizales and Ministry of Science,Technology and Innovation.

